# Extraction and Reduction of the Parameters of Archimedes Spirals Drawn by Patients

**DOI:** 10.1101/2023.04.30.23289322

**Authors:** Furrukh Khan, Jessie Xiaoxi, Brian Dalm, Evan Thomas

## Abstract

Analysis of patient’s hand drawn Archimedes spirals is commonly used in the medical community to grade various forms of tremors. These spirals are often drawn on paper using a pen or a pencil and then Xeroxed/scanned to turn the drawings into computer images. This process introduces artifacts such as misalignment of the paper, finite/variable width of the drawn line, light grey marks left by the toner, and greyscale background pixels introduced by the Xeroxing/scanning steps. Even a spiral drawn directly on the screen of a tablet produces lines with multi-pixel widths and varying greyscale values. These artifacts make it difficult to use image processing techniques to automatically extract the patient’s spiral as a clean single-valued discrete signal which could be treated mathematically for further analysis. We present a procedure in this paper to extract the patient’s hand-drawn spiral automatically as a mathematical discrete signal even in the presence of artifacts, with minimal user intervention. We also note that the spirals used by some hospitals and clinics are distorted and not perfect Archimedes spirals; nevertheless, our procedure can still be used for these cases. The extracted discrete signal is composed of a couple of thousand samples (features). The largeness of this feature space compared with the typical number of spiral samples at our disposal (of the order of only hundreds) makes it infeasible to apply Machine Learning techniques for predictions which generalize well in the real world without overfitting. We analyze the extracted discrete signal using FFT (Fast Fourier Transforms) and show that in FFT space the signal can be represented by as few as 300 parameters. The paper concludes that if these 300 parameters (or even 150 parameters for some problems) are used as a feature set for Machine Learning then it could very well be possible to make predictions which generalize well to the real world without overfitting. As a note, applications to actual Machine Learning problems are not covered in this paper.

## 1. Introduction

Discovery of the Archimedes spiral^1^ (Fig. 1) in the 3^rd^-century BC is attributed to Canon of Samos^2^ a disciple of Archimedes^3^. This spiral is extensively used in medicine to study various forms of tremors. A patient is asked to draw a spiral by hand inside the space of an Archimedes spiral and a physician grades the severity of the tremor based on the patient’s drawing. An example is shown in Fig. 2 where we have drawn the spiral ourselves to mimic a patient’s hand-drawn spiral. It is extremely beneficial to be able to convert the patient’s spiral into a mathematical function (or a signal) so that mathematics could be used to process the signal, e.g., using statistics to make inferences, or applying Machine Learning techniques to make predictions.

**Figure 1.**
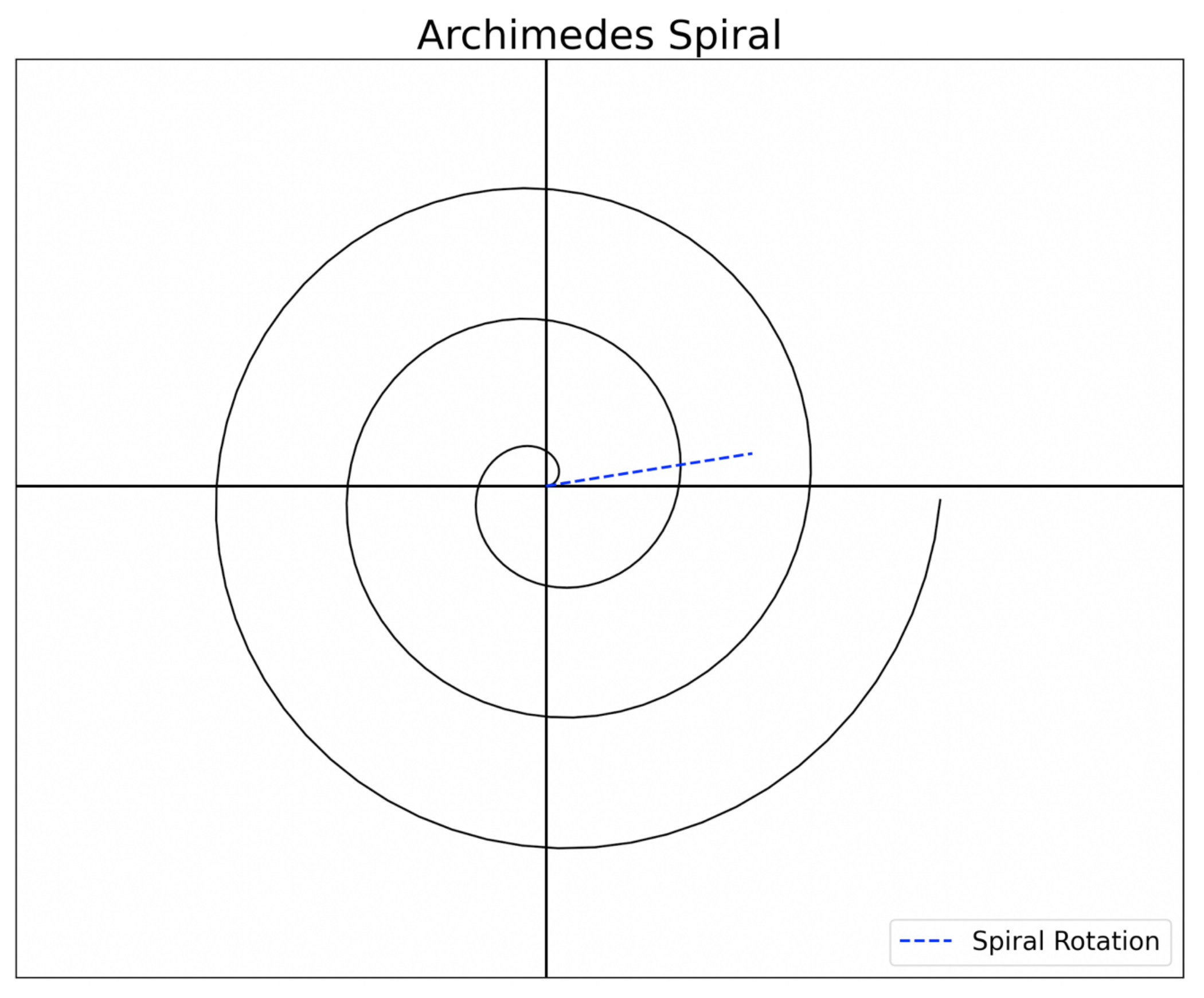
Archimedes Spiral, rotated by *θ*_*r*_ (Spiral Rotation).

**Figure 2.**
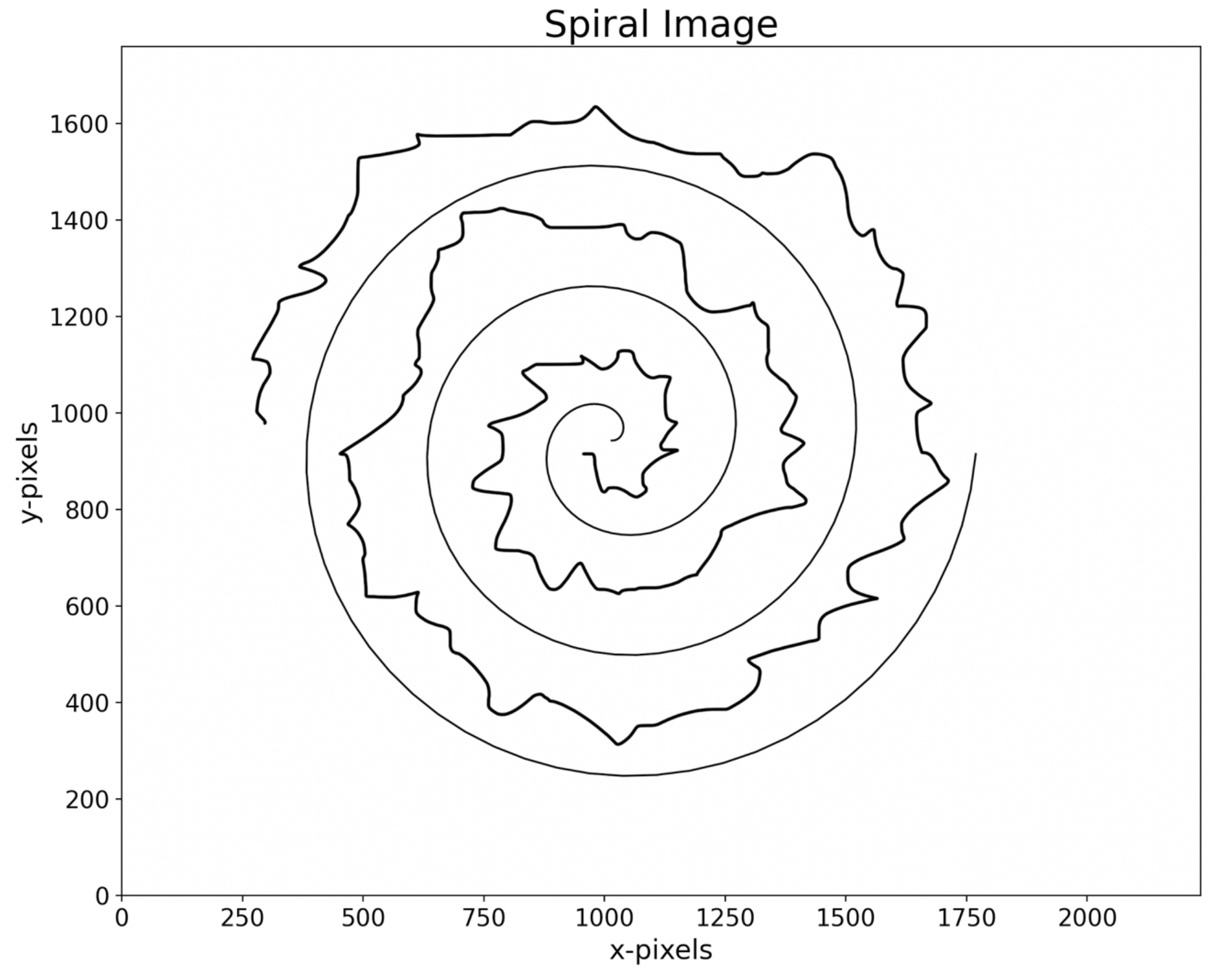
Image in *Spiral Space* showing the spiral as well as the patient’s hand-drawn spiral

The spirals are often drawn on paper by a patient using a pen or a pencil. To convert these drawings to images, that can be transferred to a computer, the paper is Xeroxed and scanned resulting in introduction of artifacts in the image. For example, the drawn lines are not single-pixel wide, they have variable multi-pixel widths (Fig. 3). The Xeroxing/scanning step can introduce regions of varying light grey pixels in the background instead of white pixels.

**Figure 3.**
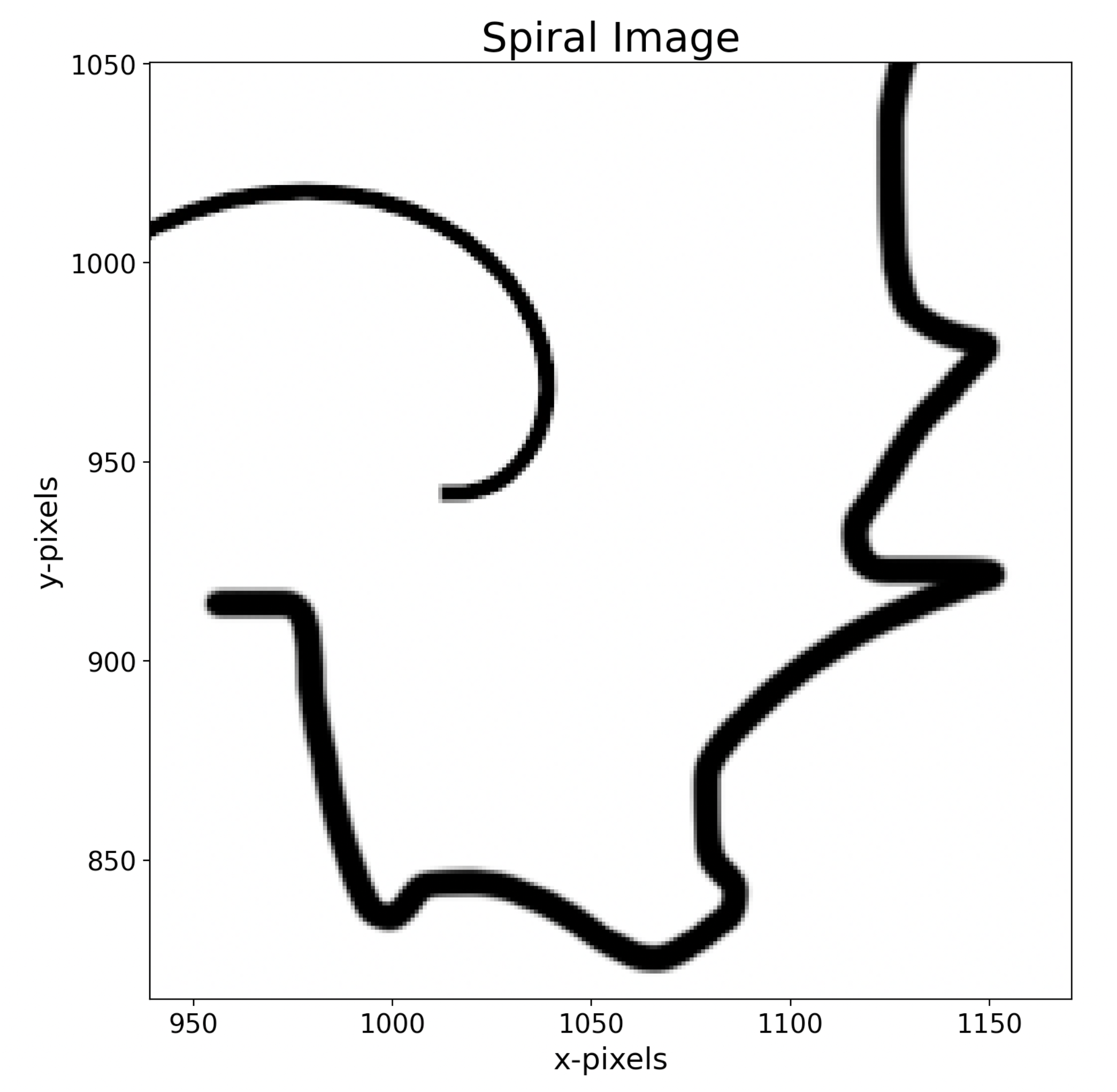
Zoomed portion of the image in Fig. 2, showing that the spiral as well as the patient’s hand-drawn spiral have multi-pixel widths with non-black greyscale values at the edges

Furthermore, documents are not placed precisely aligned with the beds of the Xerox machines/scanners which cause the spirals to get rotated and translated by unpredictable amounts.

These artifacts make it challenging to extract a clean single-valued discrete signal from the image using image processing techniques. In this paper we outline a procedure which is not based on image processing, and which automatically extracts a clean signal from an image with minimal user interaction (Fig. 8). The basic mathematics for these techniques is presented in section 2 and 3. Sections 4 and 5 explain the steps of our technique along with the software used to implement these steps.

After extracting the discrete signal (Fig. 8), this paper turns its focus on the mathematical analysis of the signal by transforming it to FFT^4^ (Fast Fourier Transform) space. Section 6 contains the steps to determine the FFT of the signal (Fig. 9), and then recover the signal back by performing an inverse FFT^5^. Section 6 shows that we can get a good approximation of the discrete signal by keeping only 151 parameters of the FFT (Fig. 10). This result has strong implications on the feasibility of using Machine Learning on spiral data to make predictions. This is explained in Section 8.

It turns out that some hospitals use spirals which are not exactly Archimedes spirals, instead they are somewhat distorted (Fig. 11). We show in section 7 that our technique needs only a small modification to be applicable to these distorted spirals as well (Fig. 17). Finally, we present the conclusions of this paper in section 8. The software to implement the entire functionality of this paper is included in Appendices A and B.

## 2. Mathematical Preliminaries

In this section we cover a few mathematical details relevant to this paper.

### 2.1 Basic Equations in Spiral Space

We start with a 2D space with polar coordinates *r*∠*θ*. We name this space *Spiral Space*. An Archimedes Spiral in this space is defined as^1^,

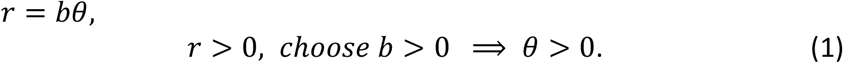

We choose *b* > 0 in this paper which implies that *θ* > 0. The spiral can be rotated around its origin by replacing *θ* by *θ*+*θ*_*r*_,

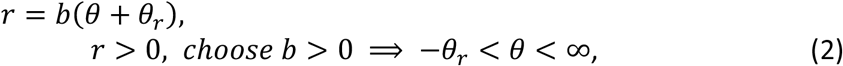

where *θ*_*r*_ is the angle of rotation. To express this rotated spiral in rectangular coordinates centered at the origin we use (2) and

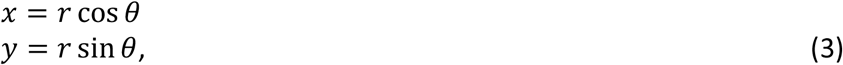

to get,

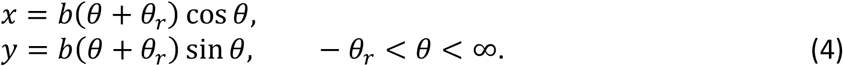

Figure 1 shows a spiral rotated by a positive angle, *θ*_*r*_.

### 2.2 Spiral equation from two points

Since eq. (4) has two unknown parameters, *b* and *θ*_*r*_, the equation for a centered spiral can be completely determined if we know the polar coordinates of any two points *r*_11_∠*θ*_11_ and *r*_22_∠*θ*_22_ lying on the spiral. Inserting the coordinates of these two points into equation (2) we get two equations in two unknowns. Solving these equations gives us the unknowns *θ*_*r*_ and *b*,

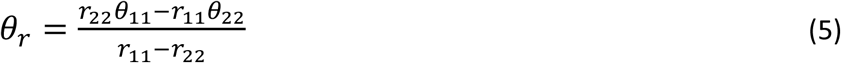

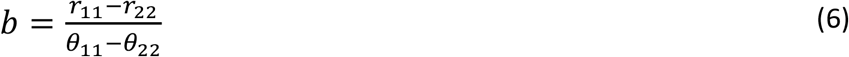

### 2.3 Flattened Space

Our aim is to map the *Spiral Space* containing the spiral expressed by (2) to a new space, which we call *Flattened Space*, in which the spiral is unraveled or flattened into a straight horizontal line. Such a mapping is given by,

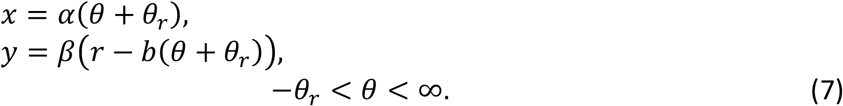

were *α* and *β* are arbitrary constants. It is easy to see by substituting (2) into (7) that we get *y* = 0 for all values of *x*. Also, as *θ* changes, *x* changes linearly with it, proportional to the value of *α*. Hence, the spiral is mapped into the *x*-axis through this mapping. In this paper we choose *α* = 1 and *β* = 1 without loss of generality.

## 3. Zones and Branch Cut in the Flattened Space

In this paper we are given an image in some format (jpg, png, tiff etc.) whose pixels (greyscale or color) contain the spiral as well as the patient’s hand-drawn spiral, Fig. 2. A zoomed in portion of this image in shown in Fig. 3. Note that the spiral and patient’s hand-drawing are not single pixel but multiple-pixel wide with different greyscale or color values. We are not given the actual equation of the spiral contained in the image pixels. To flatten the patient’s hand-drawn spiral by using (7) we need an equation for the spiral, i.e., the parameters *b* and *θ*_*r*_. Later in the paper we show explicitly how our software allows us to determine these parameters from the image, here we assume that we have access to these parameters.

As we increase *θ* in (2), *r* increases to trace the spiral, with *θ* varying from −*θ*_*r*_ to ∞. It is useful to rewrite this equation by redefining *θ* in terms of a new variable *θ*′ which is restricted to the interval 0 to 2*π*,

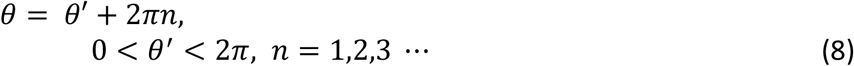

The first part of the spiral near the origin is traced by −*θ*_*r*_< *θ*′ < 2*π*.

To draw the spiral, we increase *θ*′, and every time it reaches 2*π*, i.e., the spiral crosses the negative *x-axis*, we increase *n* by 1 and reset *θ*′ to 0. We denote the negative *x-axis* as the “branch cut” in *Spiral Space*, as shown in Fig. 4.

**Figure 4.**
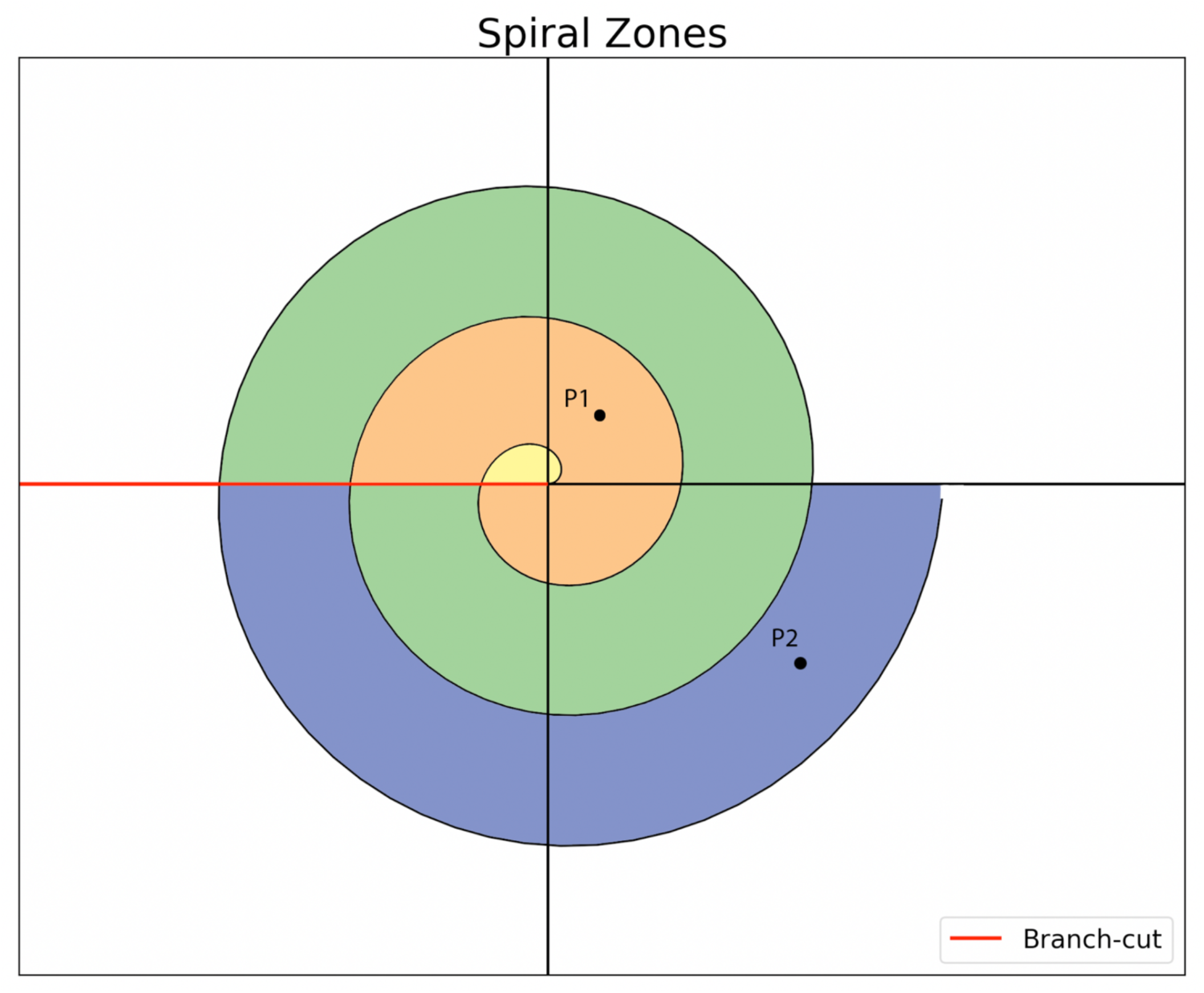
Various zones differentiated by different colors. Only part of the blue zone is shown. P1 and P2 are pixels belonging to zones with *n* = 1 and *n* = 3 respectively. Branch-cut is defined in the text.

We use (7) and (8) with *α* = *β* = 1 to map all the pixels in the image to the Flattened Space. Note that some of these pixels are form the spiral, some form the patient’s hand-drawn spiral, while most others are just white pixels (or light grey pixel artifacts introduced by the Xerox machines/scanners). Our aim is to map each pixel of the whole image, not just the spiral pixels.

This brings us to the central problem in flattening the image. If we have a random point (pixel), *r*∠*θ*, in *Spiral Space*, how do we determine which integer *n* = 1,2,3 ⋯ in (8) to assign to it so that we can use (7) and (8) to map it into *Flattened Space*? The image gives us no straightforward way to determine this number. If *n* is not determined then any point P in the *Spiral Space* will map not only to one single point (*x, y*) in the *Flattened Space* but to a family of points, each point being associated with a give *n* = 1,2,3 ⋯,

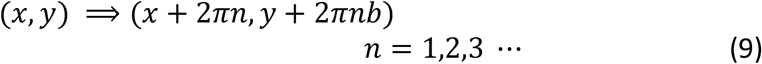

We can rephrase this problem differently by introducing the concept of zones. The set of points in *Spiral Space* which should be associated with n=1 for the mapping is assigned zone1, all points which need n=2 are grouped into zone2 etc. Fig. 4 shows some zones in *Spiral Space* in different colors. In this figure point P1 belongs to zone1 with *n* = 1, and P2 belongs to zone3 with *n* = 3. Now we rephrase the problem posed in the previous paragraph; how do we determine to which zone a random point in *Spiral Space* belongs to? It is relatively easy to visually inspect the image and assign a zone to a point, but there is no easy way to programmatically assign a random pixel from the picture to a zone. It is impractical to visually inspect and label each of the tens of thousands of pixels in the image. It is also challenge to devise image processing algorithms to automatically determine the zones, given the aforementioned artifacts present in the images.

We solve this problem by using a simple scheme with minimal user intervention. We map each point in the image to *Flattened Space* by using not just one correct value of *n*, but instead mapping that pixel to a family of points (9) in *Flattened Space* by using a list of values for *n* = 1,2,3,4. As a result, the segments of the spiral corresponding to different values of *n* automatically get stitched together (without user’s intervention) in *Flattened Space* into a family of horizontal line segments with a vertical spacings of 2*πb*, according to (9). In the same way segments of the patient’s hand-drawn spiral in different zones automatically get stitched together into a family of parallel flattened hand-drawn spirals with vertical spacings of 2*πb*. Fig. 5 shows an example of an image in *Flattened Space* using our scheme. The figure shows the family of stitched spiral segments as horizontal lines, as well as the family of stitched patient’s hand-drawn segments. Recall that *Flattened Space* is also an image containing image pixels. Our aim is to extract that part of this image which only contains the pixels of one stitched hand-drawing from the family of many parallel hand-drawings in the image. We allow the user to make this selection (crop) in our software by interactively choosing a rectangle bounding the pixels of the desired hand-drawing. This selection is saved to a new image shown in Fig. 6. Thus, this scheme allows us to solve the problem of multiple zones by employing minimal user intervention.

**Figure 5.**
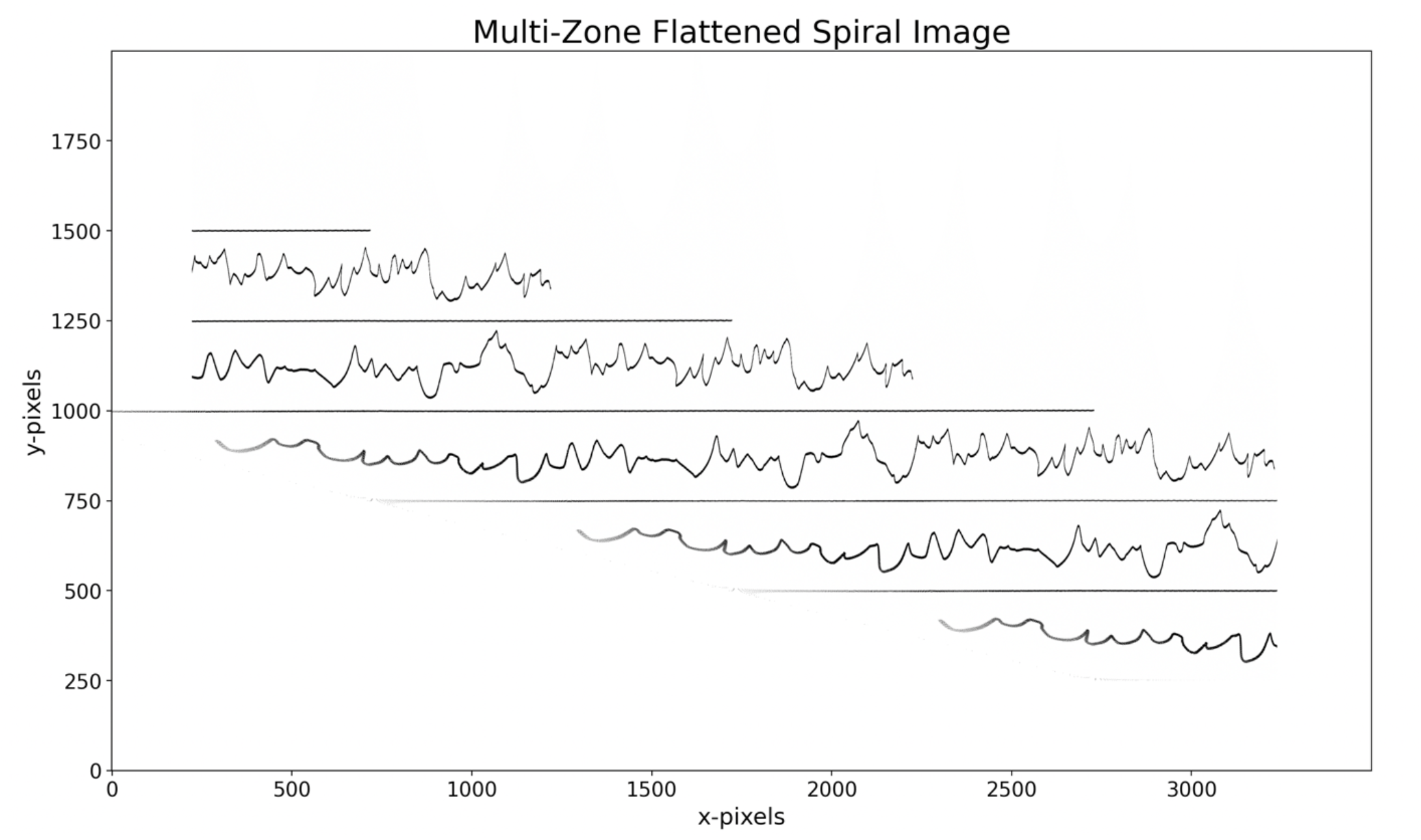
Flattened spiral image of Fig. 2 in *Flattened Space*. The horizontal lines are the family of stitched spiral segments. The wiggly lines are the family of stitched patient’s hand-drawn spiral segments.

**Figure 6.**
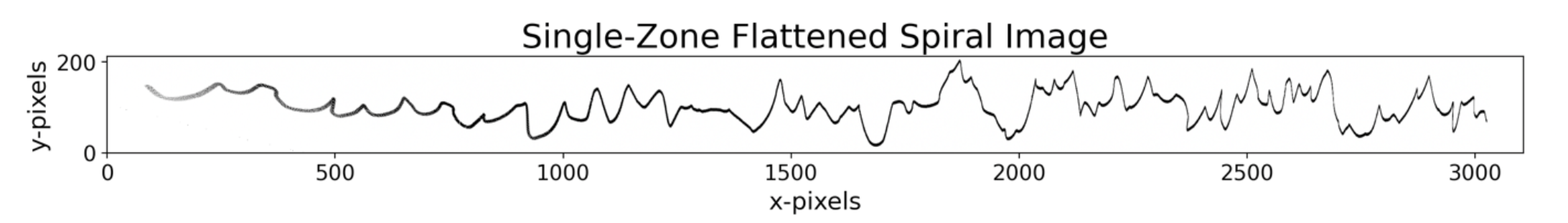
User selected pixels of a single stitched patient’s hand-drawing from the image in Fig. 5.

## 4. Software Preliminaries

All the software for this paper is included in Appendices A and B. The software is divided into steps which are explained in Sections 5 and 6. Python v3.8.2 is used to implement all the procedures presented in this paper. Python is developed under OSI (Open-Source Initiative) approved license^6,7^, making it freely usable and distributable, even for commercial use. Python’s packages *matplotlib* and *scipy* are used for plotting and scientific routines. It should be easy to convert this code to MATLAB^8^ since the Python packages used in this paper are like the corresponding implementations in MATLAB. To make the code less dependent on the unique structures of the language (Python) the code is strictly procedural, and Object Orientation is not used. This should make it easy for readers from different language backgrounds to follow the basic concepts behind the code and implement its functionality in another language such as MATLAB, C, C#, JavaScript or Java.

## 5. Obtaining the Discrete Signal from the Spiral Image

In this section we outline the steps in our approach to map the *Spiral Image* into *Flattened Space* and then extract the spatial discrete signal^9^ from the patient’s hand-drawn spiral. In the next section we outline the steps to take the DFT^10^ (Discrete Fourier Transform) of this discrete signal. We organize our approach into 5 steps. The Python program which implements these steps, Appendix A, is also arranged around these steps. This program implements interactive plots which respond to user’s mouse clicks on the screen. While a plot is waiting for user input, instructions for the user are printed on the console. We use the spiral image shown in Fig. 2 to serve as an example in the rest of this section.

### 5.1 Step 1: Read the Spiral Image file

Pixels from the image file (described in section 3) which contains the spiral as well as the patient’s hand-drawn spiral (Fig. 2) are read. Function named *mouse_press* is defined which can make a plot interactive by enabling the program to respond to user’s mouse clicks. Variable *number* counts the number of user clicks. A file which contains the log of important events is also defined.

### 5.2 Step 2: Determine the Equation of the Spiral

First, the user clicks on a plot to determine the origin of the spiral, next the user clicks twice to select two points on the spiral. The math in *section 2.2* to determine the parameters *θ*_*r*_ and *b* is followed. These parameters give us the equation of the fitted spiral (4). Fig. 7 shows that the fitted spiral is an excellent fit to the original spiral in the image file. This fitted spiral is used next to flatten the Spiral Image.

**Figure 7.**
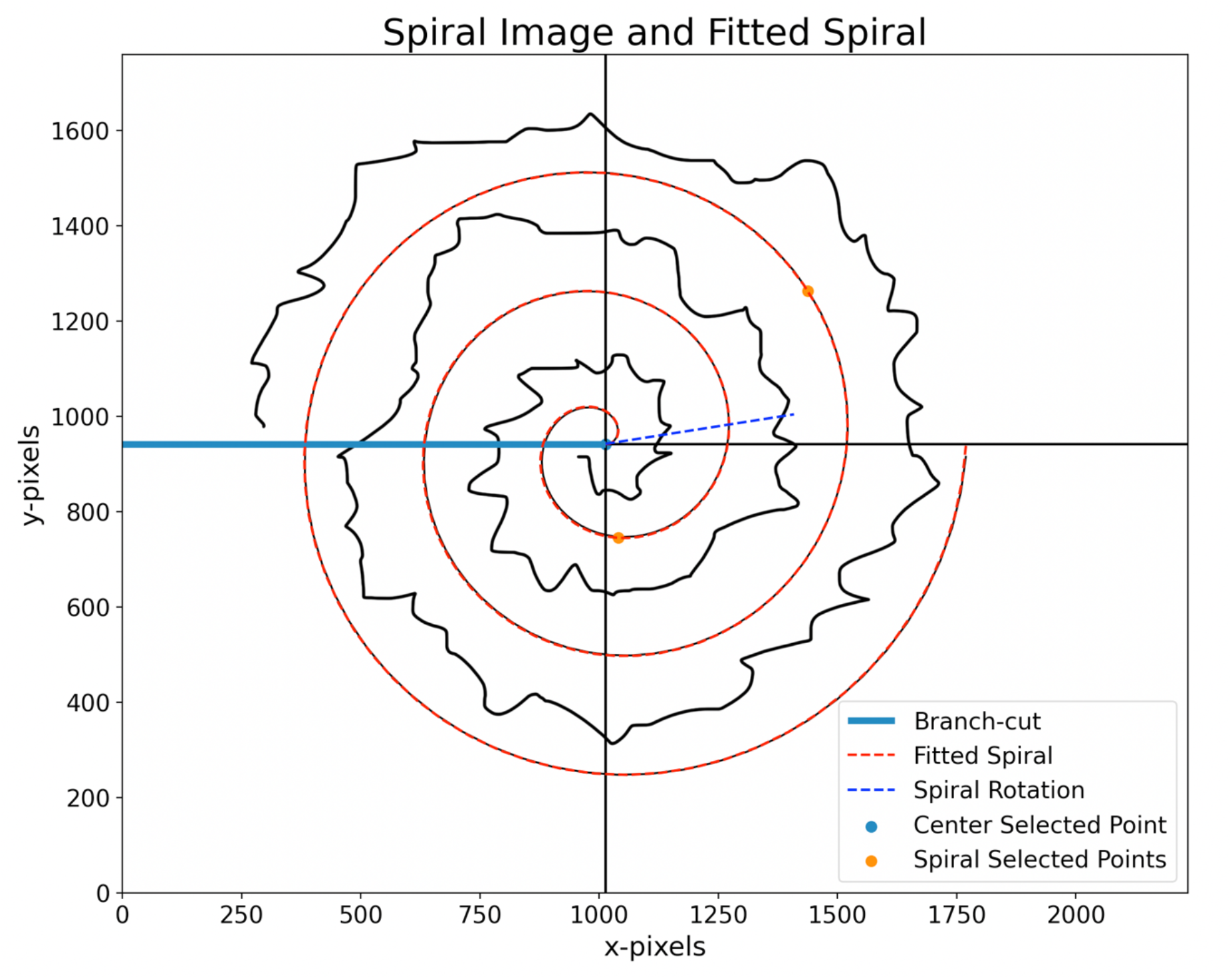
*Spiral Space* of Fig. 2 overlaid by the fitted spiral. The two points on the spiral selected by the user are also shown. The spiral is rotated by *θ*_*r*_(Spiral Rotation).

### 5.3 Step 3: Flatten the Spiral Image

Section 3 is followed and the equation for the spiral is used to map the Spiral Image to *Flattened Space*. Fig. 5 shows the family of stitched spiral segments, as well as the family of stitched patient’s hand-drawn segments. It takes about 1 min and 20 seconds on a Mac Book Pro to complete this step. This time should be far less on a fast multi-core desktop machine.

### 5.4 Step 4: Crop to get a single stitched patient’s hand-drawn spiral

The user clicks twice to select a cropping rectangle to extract the part which only contains the pixels of a single stitched patient’s hand-drawing, Fig. 6.

### 5.5 Step 5: Derive and Save the Discrete Signal

The patient’s hand-drawn spiral image from Step 5 is multiple-pixels wide with pixels having different grey scale (or color) values; it is not yet in the form of a discrete signal on which mathematical manipulations could be performed, such as determining its DFT. A further complication is provided by a scanned image having light grey (light color) pixels spread throughout the background pixels which should ideally be white. The approach we take to extract a single-valued signal is to select each x-pixel and search in the vertical direction to select the y-pixel having the highest greyscale (or color) value; we assign this y-pixel to the value of the signal corresponding to that x-pixel. A major benefit of this approach is that by choosing the highest greyscale (or color) value the background light-grey pixels present in a scanned image are completely ignored. This technique works even better if the contrast of the original image is enhanced before processing. Now we have a one-to-one correspondence between the x-values and the y-values witch constitutes a signal on which mathematical manipulations can be performed. Fig. 8 shows a signal obtained from the image which has been normalized to have zero mean and to have upper and lower bounds of ±1000. This signal can potentially have some noise spikes in a very small region near the origin; the user is prompted to crop out this noise region. The final cropped discrete signal with *N* = 2686 samples is then converted to a Panda’s data frame and saved as a csv (coma-separated value) file.

**Figure 8.**
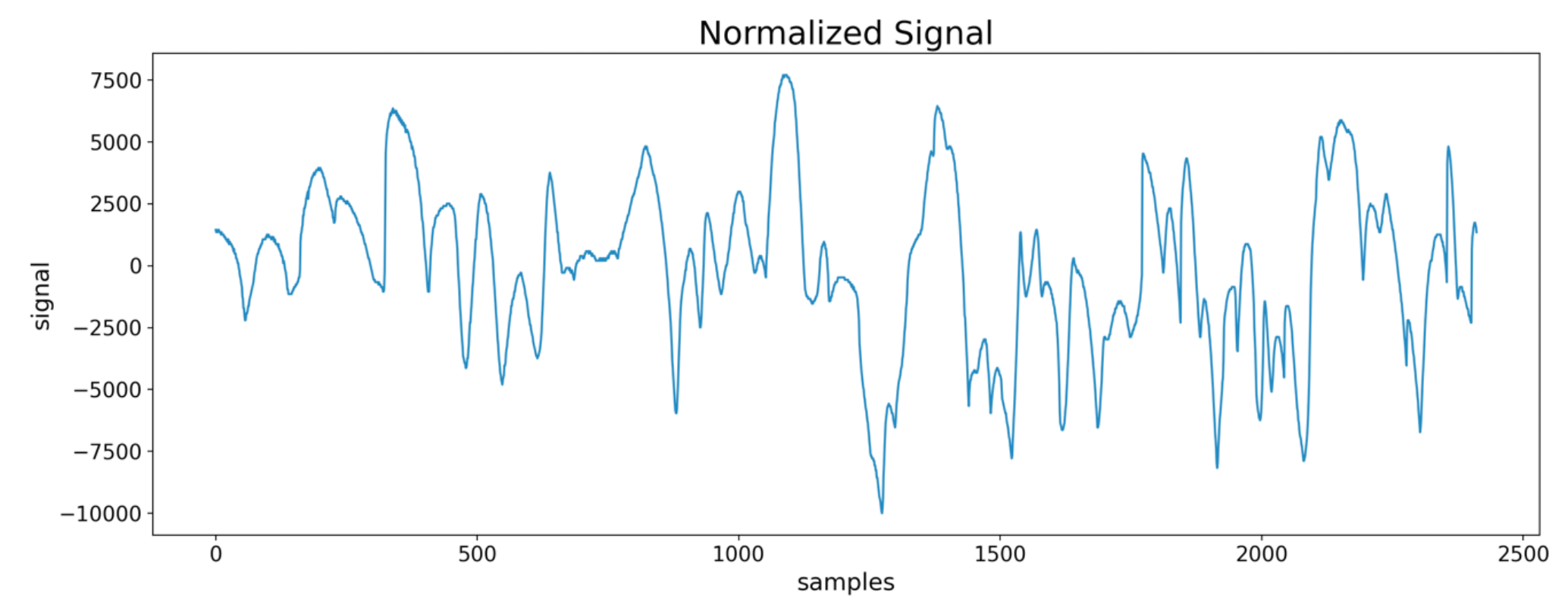
Normalized discrete signal extracted from the spiral image in Fig. 2.

## 6. Obtaining the DFT (FFT^4^) from the Signal

We organize the task of analyzing the DFT of the cropped discrete signal into 4 steps. The interactive Python program which implements these steps, Appendix B, is also partitioned into these steps. We continue to use the spiral image shown in Fig. 2 as a running example in this section also.

### 6.1 Read the Discrete Signal from the csv File

The cropped discrete signal with *N* = 2686 sample points, obtained in section 6, is read from a csv file and normalized, (normalization is explained in section 6). A file which contains the log of important events is defined.

### 6.2 Determine the FFT of the Discrete Signal

To determine the DFT, a popular and efficient algorithm named FFT (Fast Fourier Transform) is employed. The FFT of a real valued function is a complex number having a magnitude and a phase. For *N* sample points in the discrete signal, the number of coefficients, *N*_*coeff*_, in the magnitude and phase FFTs equals,

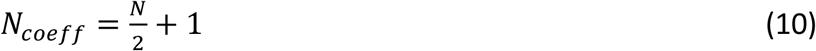

Fig. 9 shows the magnitude of the FFT obtained from the discrete signal in Fig. 8. The x-axis consists of *N*_*coeff*_ = 1344 values in this example.

**Figure 9.**
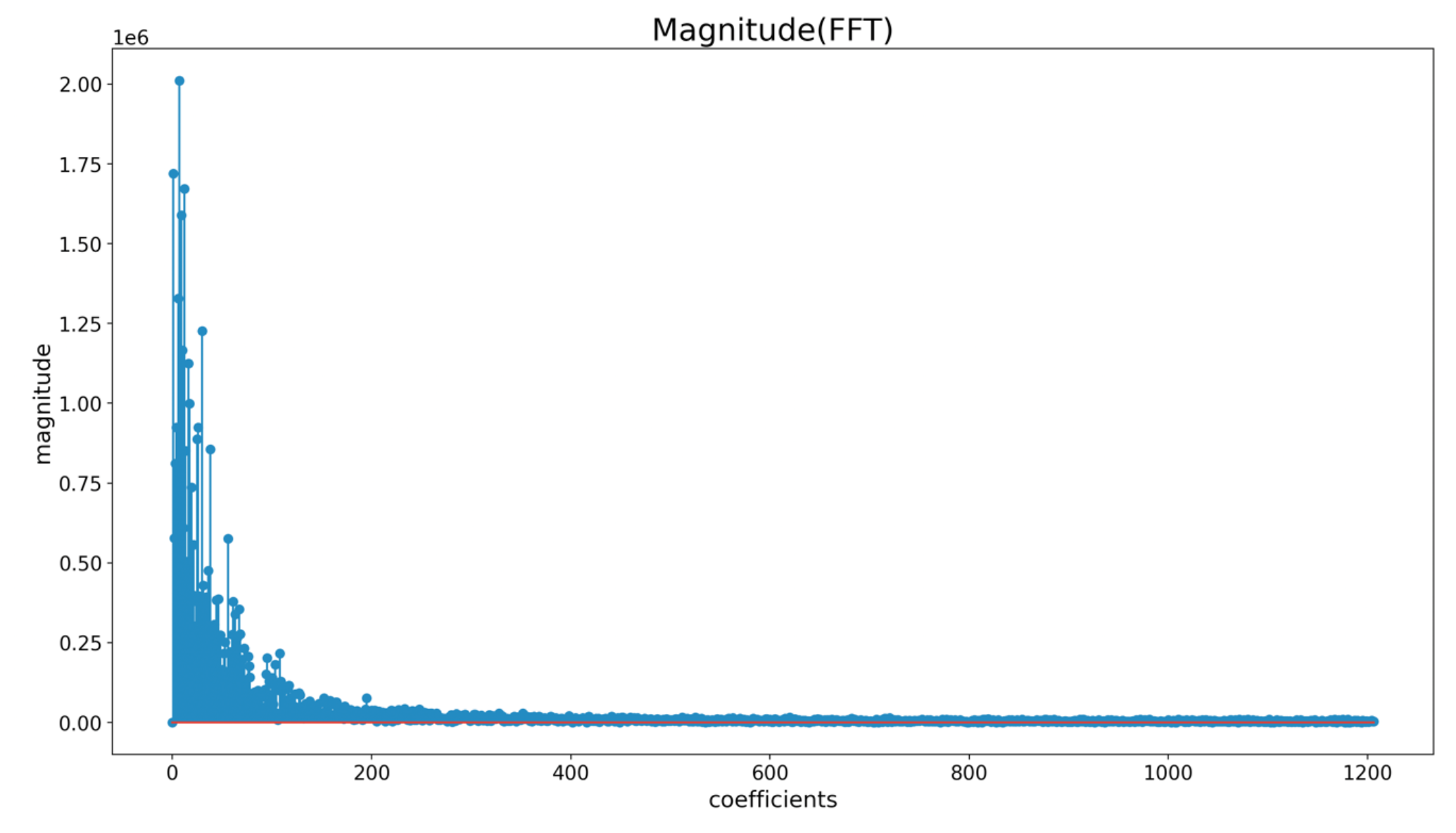
FFT of the discrete signal in Fig. 8.

### 6.3 Determine the Inverse FFT^5^ of the Truncated FFT

Fig. 9 (*N*_*coeff*_ = 1344) shows a rapid drop in the magnitude of the FFT with increasing coefficients. This observation leads us to believe that truncating the FFT at a relatively low number of coefficients, *N*_*trunc*_ ≪ *N*_*coeff*_, could result in a good approximation for the patient’s hand-drawn spiral. In other words, we may be able to approximately represent the spiral signal with only 2*N*_*trunc*_ parameters, instead of the *N* samples required for the original discrete signal. The factor of 2 in 2*N*_*trunc*_ is because we need both the magnitude and phase of the FFT to recover the discrete signal. By trying out different values of *N*_*trunc*_ on various sample spiral images we find that *N*_*trunc*_ 151 gives a good compromise between the accuracy of the discrete signal and the number of parameters required to represent it. The odd number 151 instead of 150 was chosen to get an integer value for *N* in (10).

To illustrate this on our running example, *N* = 2686 and *N*_*coeff*_ = 1344, we first truncate the FFT (both magnitude and phase) with *N*_*trunc*_ = 151 coefficients. Then we take the inverse FFT^5^ of the *truncated FFT* to get the *approximate discrete signal*. Fig. 10 shows the original discrete signal, which requires 2686 parameters, overlayed by the approximate discrete signal which requires only 2 × 151 = 302 coefficients. The figure shows that the match between the two signals is very good and for all practical purposes we can replace the original with the approximate discrete signal, which is approximately a factor of 9 reduction in the required parameters.

**Figure 10.**
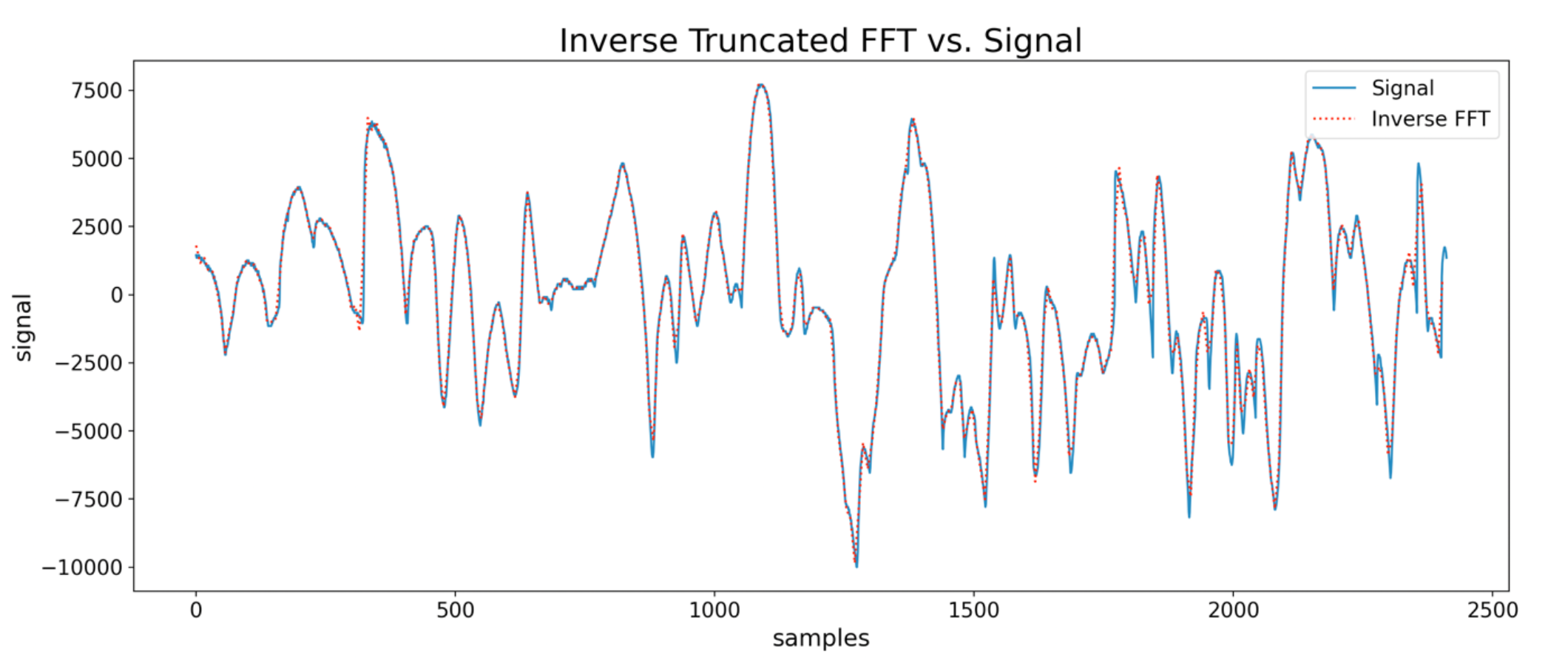
Original discrete signal overlaid by the approximate discrete signal obtained from the FFT in Fig. 9, truncated to 151 coefficients.

## 7. Distorted Archimedes Spiral

So far in this paper we have dealt with a perfect Archimedes spiral defined by (1). However, it turns out that in practice some hospitals and clinics routinely analyze patient’s hand-drawn spirals by using a particular distorted spiral which is not perfect. An example of this spiral used in actual practice is shown in Fig. 11. The patient’s hand drawing is simulated by us, but the spiral itself is a scan of a real spiral used in practice. A cursory inspection shows that it is not a perfect Archimedes spiral. We show in this section that our approach also works well on this distorted spiral simply by adding one new step in our procedure. All the steps in sections 5 and 6 remain unchanged, we simply insert a new step between steps 3 and 4.

**Figure 11.**
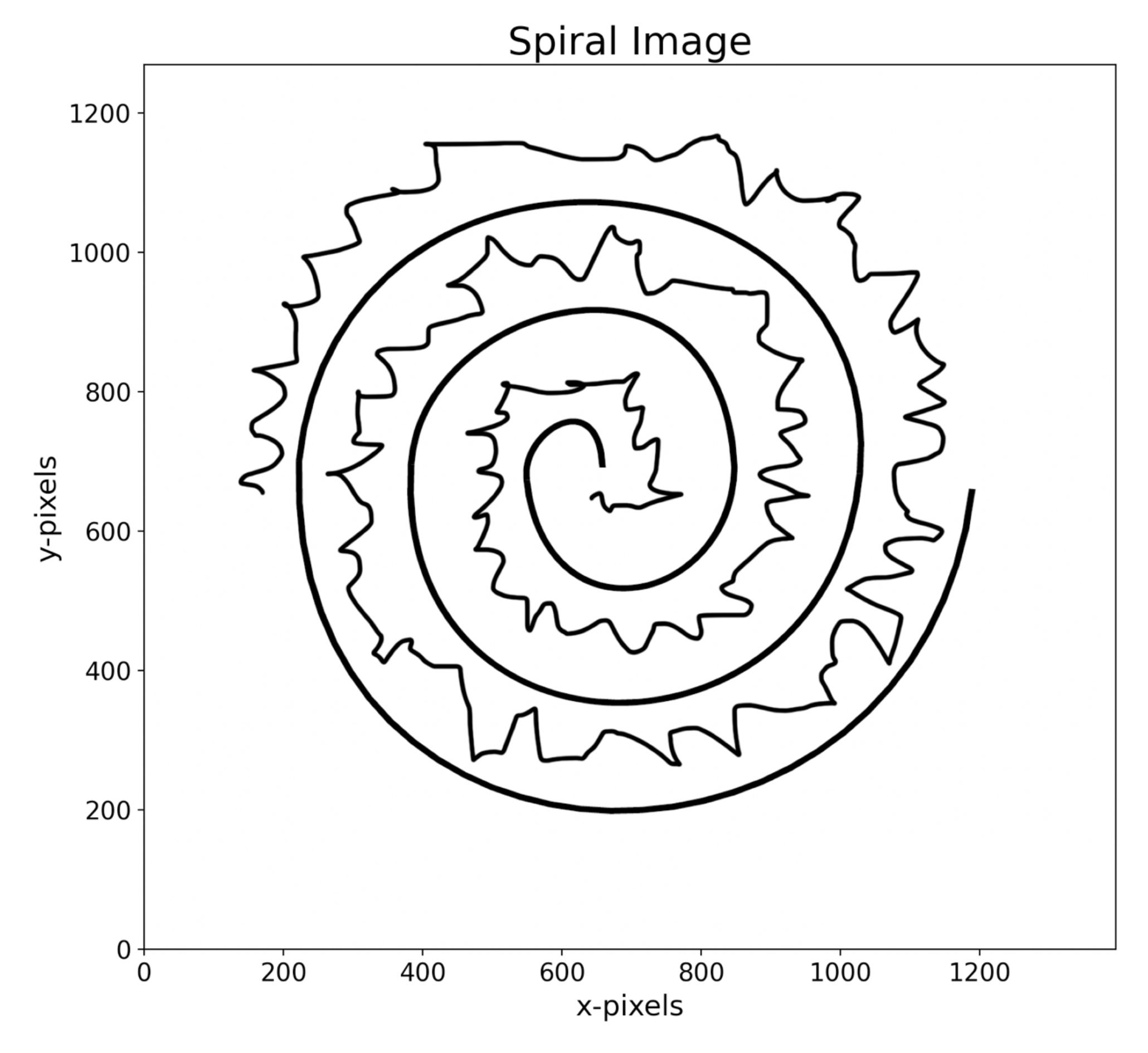
Image in *Spiral Space* showing the distorted spiral as well as the hand-drawn spiral

### 7.1 Results of Steps 1 through 3 of Section 3

We start with the distorted spiral image of Fig. 11. After selecting the center and two points on this distorted spiral we obtain an equation for the Archimedes spiral to fit the distorted spiral. The result is shown in Fig. 12. The fitted Archimedes spiral does not completely follow the distorted spiral. Flattening Fig. 11 by using the equation of the fitted Archimedes spiral gives us the family of flattened spiral images shown in Fig. 13. Since the fitted spiral in Fig. 12 does not exactly follow the distorted spiral, all the stitched spirals have a low amplitude slowly varying wave superimposed on them. For example, the distorted spiral does not map into a family of horizontal straight lines.

**Figure 12.**
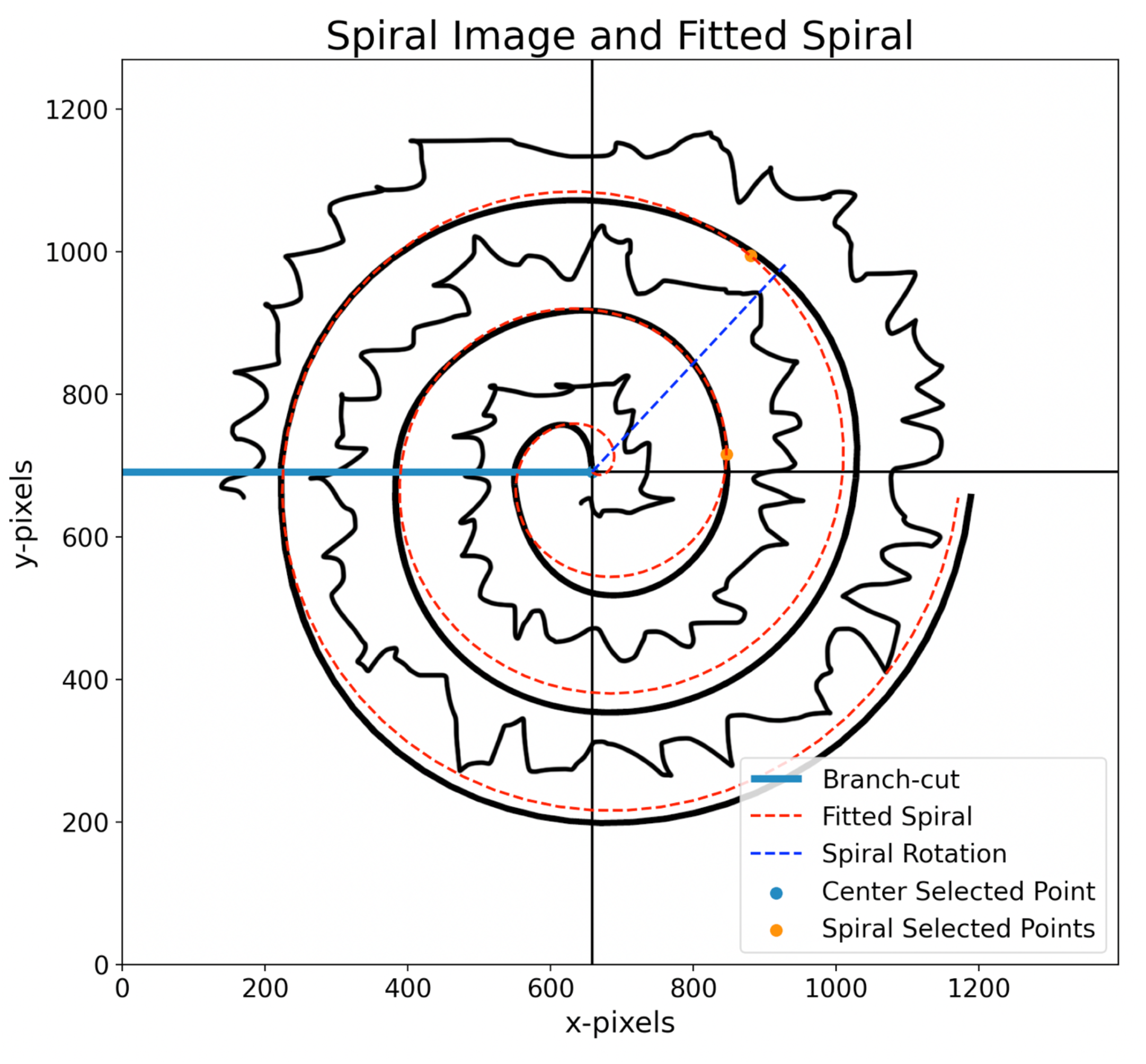
*Spiral Space* of Fig. 11 overlaid by the fitted spiral. The two points on the spiral selected by the user are also shown. The spiral is rotated by *θ*_*r*_(Spiral Rotation).

**Figure 13.**
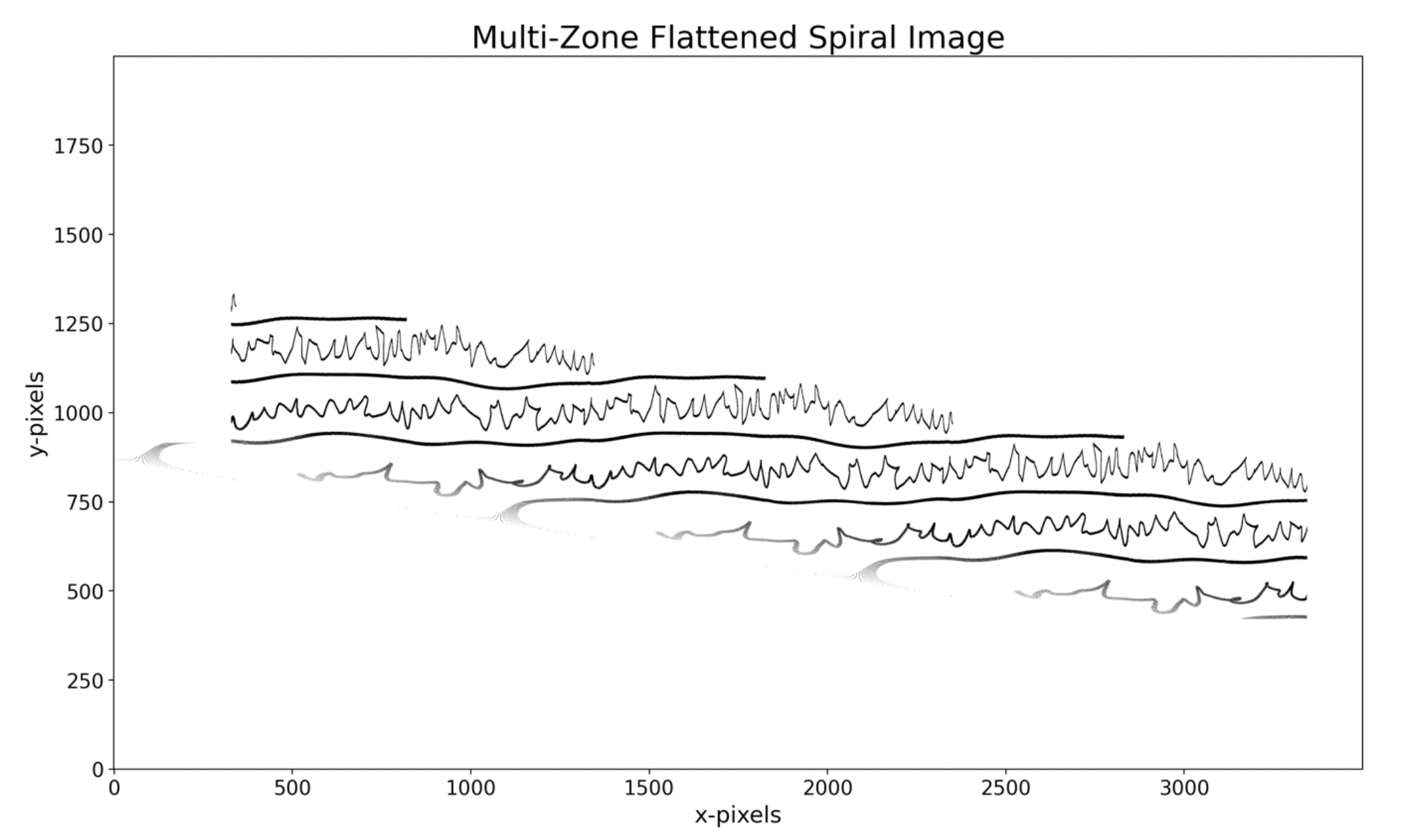
Flattened spiral Image of Fig. 11 in *Flattened Space*. The horizontal wavy lines are the family of stitched distorted spiral segments. The wiggly lines are the family of stitched patient’s hand-drawn spiral segments.

### 7.2 Step 3.5 Manually Clean the Flattened Image

The superimposed waves make it difficult to select a rectangle to cleanly crop out a single stitched patient’s spiral, because the rectangle would include pixels from other parts of the image as well. The additional step is to clean the image of Fig. 13 in an image editing application, such as Photoshop, and erase the unwanted pixels in the cropped region. The result is shown in Fig. 14. Now we can follow the rest of the steps in sections 3 and 4 without modification.

**Figure 14.**
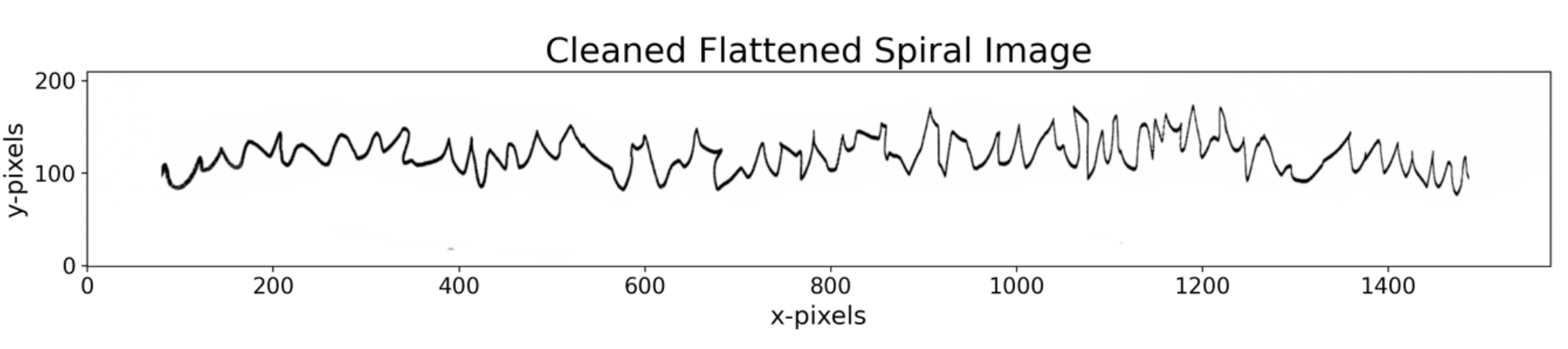
Cleaned stitched patient’s hand-drawing (in Photoshop) from the image in Fig. 13.

### 7.3 Results of Steps 4 through 5 of Section 3

By following these steps, we obtain the normalized discrete signal shown in Fig. 15.

**Figure 15.**
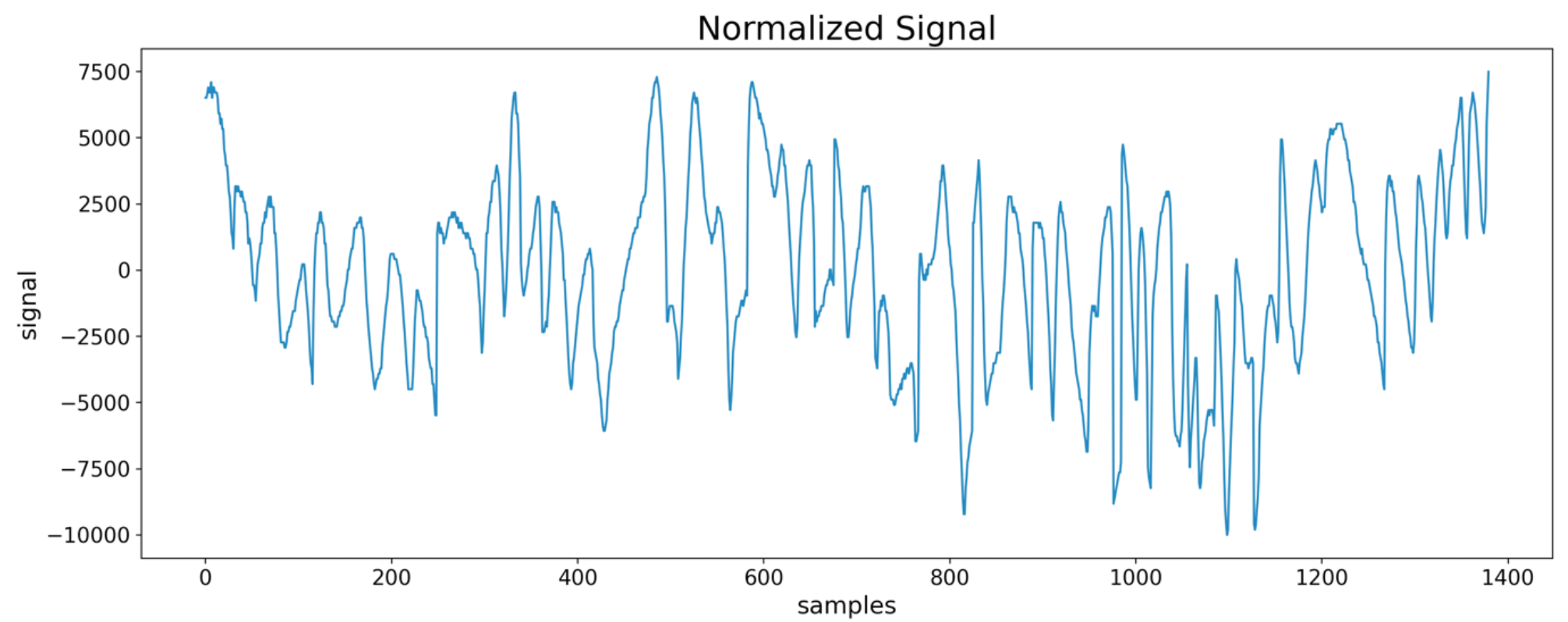
Normalized discrete signal extracted from the distorted spiral image in Fig. 11.

### 7.4 Steps 1 through 4 of Section 4

We determine the real FFT of the discrete signal and truncated the FFT by *N*_*trunc*_ = 151. The truncated magnitude FFT is shown in Fig. 15. The final approximate discrete signal is obtained via. the inverse FFT of the cropped FFT. Fig. 17 shows the original discrete signal overlayed by this approximate discrete signal. We observe that even for a distorted spiral the match between the two signals is very good and for most applications we can replace the original with the approximate discrete signal requiring only 2 × 151= 302 coefficients.

**Figure 16.**
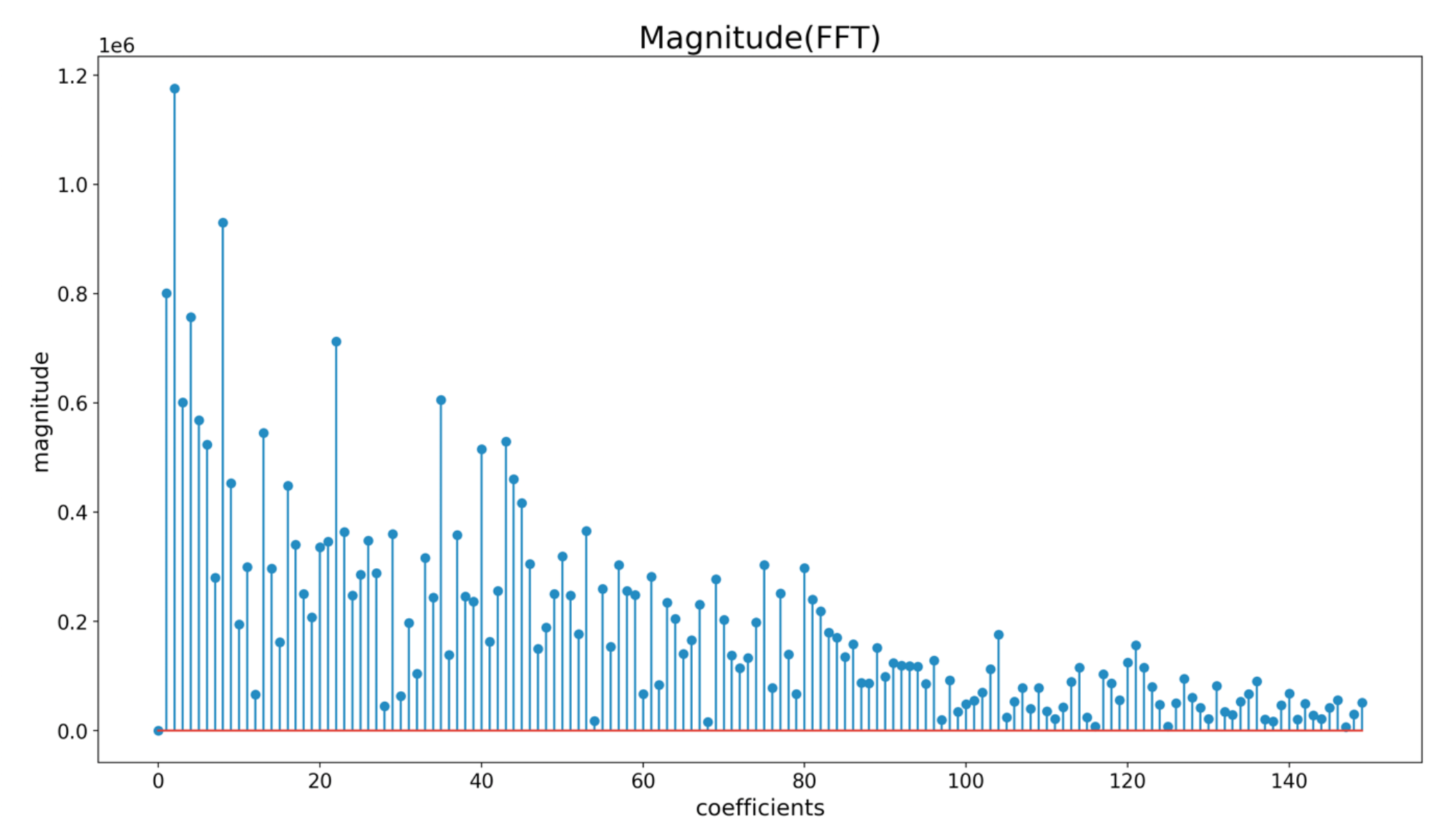
FFT of the discrete signal in Fig. 15 showing the first 151 coefficients.

**Figure 17.**
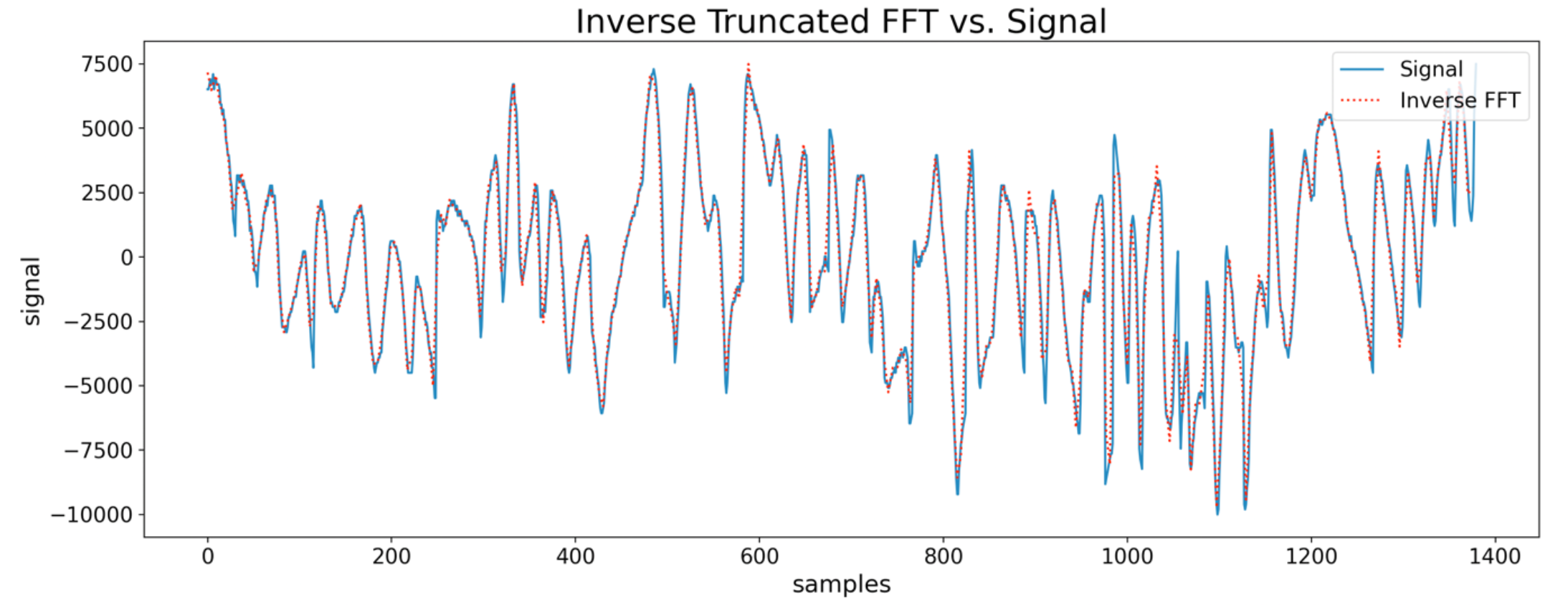
Original discrete signal from Fig. 15 overlaid by the approximate discrete signal obtained from the FFT in Fig. 16 truncated to 151 coefficients.

## 8. Conclusions

We have devised a procedure which needs minimal user intervention to automatically extract a patient’s hand-drawn spiral as a discrete signal from a scanned image, or from a drawing on a tablet’s screen. Our procedure overcomes the problem that spirals in an image are of multi-pixel width with varying pixel greyscale (or color) values, and that scans typically have light grey (or light colored) pixels in the background instead of being pure white. Furthermore, our procedure does not require precise placement of the paper (with the original image) on the scanner’s bed, it can be placed anywhere and at any angle. The fitted spiral, determined from the two points selected by the user, figures out the angle and scale of the spiral. Once the discrete signal is extracted, it can be analyzed mathematically to handle problems in various medical domains. The paper also shows that our procedure works well even for distorted spirals used by some for spiral analysis.

Additionally, we show that a signal extracted from the hand-drawn spiral can be constructed by as few as 300 coefficients, even though the signal itself could be composed of thousands of samples. This reduction of features, from thousands to a few hundred, can have important implications for applications which leverage Machine Learning to make predictions from hand-drawn spirals. For a prediction to generalize well in the real world, the amount of data (i.e., the number of hand-drawn spirals) should be at least as large as the number of features^11^ in the problem, otherwise there is real danger of overfitting^11^ and poor generalizability. Since we typically don’t have access to thousands of hand-drawn spirals, this is a serious limitation on using Machine Learning for spiral data. Using thousands of signal samples as features would potentially require thousands of hand-drawn samples. However, if we represent spirals with only 300 features then Machine Learning becomes a viable option. Furthermore, depending on the application, we may only need the magnitude FFT and not the phase, which would further reduce the features to only 150.

## Data Availability

All data produced in the present study are available upon reasonable request to the authors

## Appendix A

~~~
Appendix A
#######################################################################
# From Spiral Image to Discrete Signal
#
# STEP 1: Read the Spiral Image file
# STEP 2: Determine the equation of the spiral by fitting to image data
# STEP 3: Use the spiral equation to flatten the Spiral Image
# STEP 4: Crop to include pixels of a single stitched patient’s handdrawing
# STEP 5: Derive the discrete signal and save it to Pandas data frames
#######################################################################
from PIL import Image as im
import matplotlib.pyplot as plt
plt.rc(‘axes’, titlesize=18) 	# fontsize of the axes title
plt.rc(‘axes’, labelsize=16) 	# fontsize of the x and y labels
plt.rc(‘xtick’, labelsize=14) 	# fontsize of the tick labels
plt.rc(‘ytick’, labelsize=14) 	# fontsize of the tick labels
plt.rc(‘legend’, fontsize=14) 	# legend fontsize
import numpy as np
import math
import pandas as pd
#######################################################################
# STEP 1
# Read Spiral Image file
#######################################################################
# input filename containing the image of the spiral and the patient’s
spiral
input_file_name = ‘spiral8’ # assuming a png file
log = open(input_file_name+“_log”, ‘w’)
pr = f“input_filename = {input_file_name}”
print(pr); log.write(pr+‘\n\n’)
number = 0
coords = []
def mouse_press(event):
 global coords
 global number
 if event.inaxes:
 coords.append((event.xdata, event.ydata))
 number = number + 1
 pr = f“{number}: {event.xdata}, {event.ydata}”
 print(pr); log.write(pr+‘\n’)
# this procedure should work on png/tif/bmp
spiralImg = im.open(input_file_name + ‘.png’)
data = np.array(spiralImg)
pr = f“type(data) = {type(data) }”
print(pr); log.write(pr+‘\n’)
pr = f“y: data.shape[0] = {data.shape[0]}”
print(pr); log.write(pr+‘\n’)
pr = f“x: data.shape[1] = {data.shape[1]}”
print(pr); log.write(pr+‘\n’)
toprint = f“data.shape[2] = {data.shape[2]}”
print(pr); log.write(pr+‘\n\n’)
#######################################################################
# STEP 2
# Determine the equation of the spiral by fitting to image data
#######################################################################
pr = “--- Select the center by clicking on it ---”
print(pr); log.write(pr+‘\n’)
plt.title(‘Spiral Image’, fontsize=‘22’)
plt.xlabel(‘x-pixels’)
plt.ylabel(‘y-pixels’)
plt.imshow(np.flipud(data), origin=‘lower’)
number = 0
plt.connect(‘button_press_event’, mouse_press)
plt.show()
# select center
xcenter = coords[-1][0]
ycenter = coords[-1][1]
pr = “--- Select two points on the spiral by clicking on the points ---”
print(pr); log.write(‘\n’+pr+‘\n’)
plt.title(‘Spiral Image’, fontsize=‘22’)
plt.xlabel(‘x-pixels’)
plt.ylabel(‘y-pixels’)
plt.imshow(np.flipud(data), origin=‘lower’)
plt.connect(‘button_press_event’, mouse_press)
number = 0
plt.axvline(x=xcenter, c=“skyblue”, label=“x=0”)
plt.axhline(y=ycenter, c=“skyblue”, label=“y=0”)
plt.show()
# select 2 points of the “scanned” spiral
x11 = coords[-2][0] - xcenter
y11 = coords[-2][1] - ycenter
x22 = coords[-1][0] - xcenter
y22 = coords[-1][1] - ycenter
# determine parameters of the spiral fitted to the two points
r11 = math.sqrt(x11*x11 + y11*y11)
r22 = math.sqrt(x22*x22 + y22*y22)
t11 = np.arctan2(y11,x11)
t22 = np.arctan2(y22,x22) + 2*np.pi
t_rr = (r22*t11 - r11*t22)/(r11-r22)
t_rr = t_rr - 2*np.pi
bb = (r11-r22)/(t11-t22)
pr = f“t_rr = {t_rr} radians --- {t_rr*180.0/np.pi} degrees”
print(pr); log.write(pr+‘\n’)
pr = f“bb = {bb}”
print(pr); log.write(pr+‘\n’)
pr = “--- Close this window and wait for the next window … It will take
about 1 min and 20 sec. for this window to close …”
print(pr); log.write(‘\n’+pr+‘\n\n’)
pr = “… and the next window to appear ---”
print(pr); log.write(pr+‘\n’)
# plot fitted spiral along with the original scanned spiral
plt.gca().set_aspect(‘equal’)
# plt.connect(‘button_press_event’, mouse_press)
plt.title(‘Spiral Image and Fitted Spiral’, fontsize=‘22’)
plt.xlabel(‘x-pixels’)
plt.ylabel(‘y-pixels’)
plt.imshow(np.flipud(data), origin=‘lower’)
plt.axvline(x=xcenter, c=“black”)
plt.axhline(y=ycenter, c=“black”)
plt.hlines(y=ycenter, xmin=0, xmax=xcenter, linewidth=4, label=“Branch-cut”)
# construct and plot the fitted spiral passing through the two points
t = np.arange(−1*t_rr, 6*np.pi, 0.1)
xf=bb*(t+t_rr)*np.cos(t) + xcenter
yf=bb*(t+t_rr)*np.sin(t) + ycenter
plt.plot(xf,yf, linestyle = ‘dashed’, c=“red”, label=“Fitted Spiral”)
x_rotation = [0+xcenter, 400*np.cos(t_rr)+xcenter]
y_rotation = [0+ycenter, 400*np.sin(t_rr)+ycenter]
plt.plot(x_rotation,y_rotation, linestyle = ‘dashed’, c=“blue”,
label=“Spiral Rotation”)
# plot the centter and two points throough which the fitted spiral passes
# plt.scatter([0+xcenter, x11+xcenter, x22+xcenter], [0+ycenter,
y11+ycenter, y22+ycenter], marker = ‘o’)
plt.scatter([0+xcenter], [0+ycenter], marker = ‘o’, label=“Center Selected
Point”)
plt.scatter([x11+xcenter, x22+xcenter], [y11+ycenter, y22+ycenter], marker
= ‘o’, label=“Spiral Selected Points”)
plt.legend(loc=‘lower right’)
plt.show()
#######################################################################
# STEP 3
# Use the spiral equation ot flatten the Spiral Image
#######################################################################
# transformation to unravel the spiiral r = b*(t+t_r)
def trans(r, t, b, t_r):
 return [t + t_r, r - b*(t + t_r)]
# factor = 2*pi*n
def factor(r, t, b, t_r):
 factor = 0
 if( r > 0 and r <= b*(t + t_r)):
 # print(“zone one”)
 factor = -2*math.pi
 elif (r > b*(t + t_r) and r <= b*(t + t_r) + 2*math.pi):
 # print(“zone two”)
 factor = 0*math.pi
 elif (r > b*(t + t_r + 2*math.pi) and r <= b*(t + t_r) + 4*math.pi):
 # print(“zone three”)
 factor = 2*math.pi
 elif (r > b*(t + t_r + 4*math.pi) and r <= b*(t + t_r) + 6*math.pi):
 # print(“zone four”)
 factor = 4*math.pi
 elif (r > b*(t + t_r + 6*math.pi) and r <= b*(t + t_r) + 8*math.pi):
 # print(“zone five”)
 factor = 6*math.pi
 elif (r > b*(t + t_r + 8*math.pi) and r <= b*(t + t_r) + 10*math.pi):
 # print(“zone six”)
 factor = 8*math.pi
 return factor
# transform (flatten) two points on the spiral
r11 = math.sqrt(x11*x11 + y11*y11)
t11 = math.atan2(y11, x11) + 2*np.pi
r22 = math.sqrt(x22*x22 + y22*y22)
t22 = math.atan2(y22, x22) + 4*np.pi
# transform point 1
p11 = trans(r11, t11, bb, t_rr)
px11 = p11[0]
py11 = p11[1]
# transform point 2
p22 = trans(r22, t22, bb, t_rr)
px22 = p22[0]
py22 = p22[1]
rdata = np.full((2000, 3500, 3), [255, 255, 255])
xshift = 700
yshift = 750
for i in range(data.shape[1]): # x
 for j in range(data.shape[0]): # y
 # data[i][j] is a 1d array with 3 or 4 elements
 # each element can be accessed as data[i][j][0] etc.
 x = i - xcenter
 y = j - ycenter
 r = math.sqrt(x*x + y*y)
 t = math.atan2(y, x)
 t = t + factor(r, t, bb, t_rr)
 d = trans(r, t, bb, t_rr)
 x = d[0]
 y = d[1]
 ii = int(x*4000/25) + xshift
 jj = int(y) + yshift
 if (((rdata.shape[0] - 1)-jj) >=0 and ((rdata.shape[0] - 1)-jj) <
rdata.shape[0]) and (ii >=0 and ii < rdata.shape[1]) :
 rdata[(rdata.shape[0] - 1)-jj][ii] = data[(data.shape[0]-1)-
j][i]
for i in range(data.shape[1]): # x
 for j in range(data.shape[0]): # y
 # data[i][j] is a 1d array with 3 or 4 elements
 # each element can be accessed as data[i][j][0] etc.
 x = i - xcenter
 y = j - ycenter
 r = math.sqrt(x*x + y*y)
 t = math.atan2(y, x)
 t = t + factor(r, t, bb, t_rr)
 d = trans(r, t, bb, t_rr)
 x = d[0]
 y = d[1]
 ii = int((x+2*np.pi)*4000/25) + xshift
 jj = int(y - bb*2*np.pi) + yshift
 if (((rdata.shape[0] - 1)-jj) >=0 and ((rdata.shape[0] - 1)-jj) <
rdata.shape[0]) and (ii >=0 and ii < rdata.shape[1]) :
 rdata[(rdata.shape[0] - 1)-jj][ii] = data[(data.shape[0]-1)-
j][i]
for i in range(data.shape[1]): # x
 for j in range(data.shape[0]): # y
 # data[i][j] is a 1d array with 3 or 4 elements
 # each element can be accessed as data[i][j][0] etc.
 x = i - xcenter
 y = j - ycenter
 r = math.sqrt(x*x + y*y)
 t = math.atan2(y, x)
 t = t + factor(r, t, bb, t_rr)
 d = trans(r, t, bb, t_rr)
 x = d[0]
 y = d[1]
 ii = int((x+4*np.pi)*4000/25) + xshift
 jj = int(y - bb*4*np.pi) + yshift
 if (((rdata.shape[0] - 1)-jj) >=0 and ((rdata.shape[0] - 1)-jj) <
rdata.shape[0]) and (ii >=0 and ii < rdata.shape[1]) :
 rdata[(rdata.shape[0] - 1)-jj][ii] = data[(data.shape[0]-1)-
j][i]
pr = “--- Select two points on the graph to define a rectangle. Only data
inside this rectangle will be kept ---”
print(pr); log.write(pr+‘\n’)
plt.title(‘Multi-Zone Flattened Spiral Image’, fontsize=‘22’)
plt.xlabel(‘x-pixels’)
plt.ylabel(‘y-pixels’)
plt.imshow(np.flipud(rdata), origin=‘lower’)
number = 0
plt.connect(‘button_press_event’, mouse_press)
plt.show()
#######################################################################
# STEP 4
# Crop to include pixels of a single stitched patient, Äôs hand-drawing
#######################################################################
# select 2 points for the selection to choose data
xselection_up_left = int(coords[-2][0])
yselection_up_left = int(coords[-2][1])
xselection_down_right= int(coords[-1][0])
yselection_down_right = int(coords[-1][1])
hselection = yselection_up_left - yselection_down_right
wselection = xselection_down_right - xselection_up_left
sdata = np.full((hselection, wselection, 3), [255, 255, 255])
for i in range(xselection_up_left, xselection_up_left + sdata.shape[1]): #
x
 for j in range(yselection_down_right, yselection_down_right +
sdata.shape[0]): # y
 ii = i - xselection_up_left
 jj = j -yselection_down_right
sdata[(sdata.shape[0] - 1)-jj][ii] = rdata[(rdata.shape[0]-1)-
j][i]	
pr = “--- Select two points on the graph to define a rectangle. Picture
data inside this rectangle will be converted to a discrete signal. … ---
print(pr); log.write(‘\n’+pr+‘\n’)
plt.imshow(np.flipud(sdata), origin=‘lower’)
plt.title(‘Single-Zone Flattened Spiral Image’, fontsize=‘22’)
plt.xlabel(‘x-pixels’)
plt.ylabel(‘y-pixels’)
number = 0
plt.connect(‘button_press_event’, mouse_press)
plt.show()
#######################################################################
# STEP 5
# Derive the discrete signal and save it to Pandas data frames
#######################################################################
xselection_up_left = int(coords[-2][0])
yselection_up_left = int(coords[-2][1])
xselection_down_right= int(coords[-1][0])
yselection_down_right = int(coords[-1][1])
# derive discrete signal disc_signal[i], i = 0, 1, 2, 3 … from sdata
disc_signal = []
for i in range(xselection_up_left, xselection_down_right):
 min = 300*3
 jmin = 0
 for j in range(yselection_down_right, yselection_up_left):
 if (sdata[(sdata.shape[0] - 1)-j][i][0]+sdata[(sdata.shape[0] -
1)-j][i][1]+sdata[(sdata.shape[0] - 1)-j][i][2] < min):
 min = sdata[(sdata.shape[0] - 1)-
j][i][0]+sdata[(sdata.shape[0] - 1)-j][i][1]+sdata[(sdata.shape[0] - 1)-
j][i][2]
 jmin = (sdata.shape[0] - 1)-j
 disc_signal.append(jmin)
pr = “--- Depending on the previous selection, this discrete signal likely
contains noise spikes on one or both sides …”
print(pr); log.write(‘\n’+pr+‘\n’)
pr = “… Select two points on the graph to define an x-interval which
excludes noise in the discrete signal ---”
print(pr); log.write(pr+‘\n’)
plt.plot(disc_signal)
plt.title(‘Uncropped Discrete Signal’, fontsize=‘22’)
plt.xlabel(‘samle’)
plt.ylabel(‘signal’)
number = 0
plt.connect(‘button_press_event’, mouse_press)
plt.show()
# convert disc_signal to pandas data frame and save to disc
dict = {input_file_name+‘_uncropped_signal’: disc_signal}
df = pd.DataFrame(dict)
df.to_csv(input_file_name+‘_disc_uncropped_signal.csv’, index=False)
pr = f“--- ‘discrete uncropped signal’ has been saved as
{input_file_name+‘_disc_uncropped_signal.csv’} ---”
print(pr); log.write(‘\n’+pr+‘\n’)
# select 2 points to crop to a narrower selection to choose signal data
without noise
pr = “--- This is the final discrete signal ---”
print(pr); log.write(‘\n’+pr+‘\n’)
xselection_left = int(coords[-2][0])
xselection_right= int(coords[-1][0])
width_selection = xselection_right - xselection_left
final_disc_signal = []
for i in range(xselection_left, xselection_right):
 final_disc_signal.append(disc_signal[i])
plt.title(‘Final Discrete Signal’, fontsize=‘22’)
plt.xlabel(‘sample’)
plt.ylabel(‘signal’)
plt.plot(final_disc_signal)
plt.show()
# convert final_disc_signal to pandas data frame and save to disc
dict = {input_file_name+‘_signal’: final_disc_signal}
df = pd.DataFrame(dict)
df.to_csv(input_file_name+‘_disc_signal.csv’, index=False)
pr = f“--- ‘final discrete signal’ has been saved as
{input_file_name+‘_disc_signal.csv’} ---”
print(pr); log.write(‘\n’+pr+‘\n’)
log.close()
~~~

## Appendix B

~~~
#######################################################################
# From Discrete Signal to FFT
#
# STEP 1: Read Discrete Signal from csv file
# STEP 2: Determine the Real FFT of the Discrete Signal
# STEP 3: Determine the Inverse FFT of the truncated FFT
# STEP 4: Save the coefficients of the truncated FFT to Pandas data frames
#
#######################################################################
import matplotlib.pyplot as plt
plt.rc(‘axes’, titlesize=18) # fontsize of the axes title
plt.rc(‘axes’, labelsize=16) # fontsize of the x and y labels
plt.rc(‘xtick’, labelsize=14) # fontsize of the tick labels
plt.rc(‘ytick’, labelsize=14) # fontsize of the tick labels
plt.rc(‘legend’, fontsize=14) # legend fontsize
import numpy as np
import pandas as pd
from scipy.fft import rfft, rfftfreq
from scipy.fft import irfft
#######################################################################
# STEP 1
# Read Discrete Signal from csv file
#######################################################################
# input csv filename containing the discrete signal
input_file_name = ‘spiral8_disc_signal’
# input_file_name = ‘Spiral10_flattened4_disc_signal’
# Choose first N_FFT_truncate coefficients of the spectrum
N_FFT_truncate = 151
log = open(input_file_name+“_log”, ‘w’)
pr = f“input_filename = {input_file_name}”
print(pr); log.write(pr+‘\n\n’)
# Read signal from the csv file
df1 = pd.read_csv(input_file_name + “.csv”)
# Need to flatten to change shape froom (n, 1) to (n,)
df2 = df1.to_numpy().flatten()
# subtract the dc value
aver = np.average(df2)
df3 = df2 - aver
# Normalize
df = np.int16((df3 / np.abs(df3).max()) * 10000)
plt.plot(df)
plt.title(‘Normalized Signal’, fontsize=‘22’)
plt.xlabel(‘samples’)
plt.ylabel(‘signal’)
plt.show()]
#######################################################################
# STEP 2
# Determine the Real FFT of the Discrete Signal
#######################################################################
# rfft stands for real fft
# Number of sample points, N
N = df.shape[0]
pr = f“Number of sample points: N = {N}”
print(pr); log.write(pr+‘\n\n’)
yf = rfft(df)
# xf contains the frequencies
xf = rfftfreq(N, 1)
# Number of coefficients in rfft will equal N/2 + 1
pr = f“Number of coefficients in the transform, N/2 + 1:
{np.shape(yf)[0]}”
print(pr); log.write(pr+‘\n’)
plt.stem(np.abs(yf))
plt.title(‘Magnitude(FFT)’, fontsize=‘22’)
plt.xlabel(‘coefficients’)
plt.ylabel(‘magnitude’)
plt.show()
# inverse rfft using irfft
iyf = irfft(yf)
plt.plot(df, label=“Signal”, c=“blue”)
plt.plot(iyf, linestyle = ‘dashed’, c=“red”, linewidth=“2”, label=“Inverse
FFT”)
plt.title(‘Inverse FFT vs. Signal’, fontsize=‘22’)
plt.xlabel(‘samples’)
plt.ylabel(‘signal’)
plt.legend(loc=‘upper right’)
plt.show()
#######################################################################
# STEP 3
# Determine the Inverse FFT of the truncated FFT
#######################################################################
pr = f“Number of coefficients in the transform after truncation:
{N_FFT_truncate}”
print(pr); log.write(pr+‘\n’)
yf_trunc = []
for i in range(N_FFT_truncate):
 yf_trunc.append(yf[i])
plt.stem(np.abs(yf_trunc))
plt.title(‘Magnitude(FFT)’, fontsize=‘22’)
plt.xlabel(‘coefficients’)
plt.ylabel(‘magnitude’)
plt.show()
plt.stem(np.angle(yf_trunc))
plt.title(‘Phase(FFT)’, fontsize=‘22’)
plt.xlabel(‘coefficients’)
plt.ylabel(‘phase’)
plt.show()
# Use irfft to determine the inverse transform from the truncated spectrum
N_inv = (N_FFT_truncate-1)*2
iyf_trunc = irfft(yf_trunc, n=N_inv)
scale = N/N_inv
ixf = []
for i in range(iyf_trunc.shape[0]):
 ixf.append(int(i*scale))
# Plot the inverse transform from the truncated spectrum on top of
original signal
plt.plot(df, label=‘Signal’, c=‘blue’)
# plt.plot(ixf, iyf_trunc/scale, linestyle = ‘dotted’, marker=‘.’,
c=“red”, label=‘Inverse FFT’)
plt.plot(ixf, iyf_trunc/scale, linestyle = ‘dashed’, c=“red”,
label=‘Inverse FFT’, linewidth=“2”)
# plt.plot(ixf, iyf_trunc/scale, c=“red”, label=‘Inverse FFT’)
plt.title(‘Inverse Truncated FFT vs. Signal’, fontsize=‘22’)
plt.xlabel(‘samples’)
plt.ylabel(‘signal’)
plt.legend(loc=‘upper right’)
plt.show()
#######################################################################
# STEP 4
# Save the coefficients of the truncated FFT to Pandas data frames
#######################################################################
# Save cropped coeffiecien to pandas data frame and save to disc
dict = {‘Magnitude’: np.abs(yf_trunc), “Phase”: np.angle(yf_trunc)}
df = pd.DataFrame(dict)
df.to_csv(input_file_name+‘_trunc_spectrum.csv’, index=False)
pr = f“Truncated spectrum has been saved as
{input_file_name+‘_trunc_spectrum.csv’}”
print(pr); log.write(‘\n’+pr+‘\n’)
df2 = df.drop([‘Phase’], axis = 1)
df2 = df2.T
column_names = []
for i in range(N_FFT_truncate):
 column_names.append(“m”+str(i))
df2.columns = column_names
df3 = df.drop([‘Magnitude’], axis = 1)
df3 = df3.T
column_names = []
for i in range(N_FFT_truncate):
 column_names.append(“p”+str(i))
df3.columns = column_names
df4 = pd.concat([df2.reset_index(drop=True), df3.reset_index(drop=True)],
axis=1)
df4.to_csv(input_file_name+‘_trunc_spectrum_features.csv’, index=False)
pr = f“Transpose of truncated spectrum as features has been saved as
{input_file_name+‘_trunc_spectrum_feaatures.csv’}”
print(pr); log.write(pr+‘\n’)
log.close()
~~~

